# Climate Change and Child Health - Assessment of Parents’ Perspective and Relevance

**DOI:** 10.1101/2021.02.15.21251730

**Authors:** Lena Lagally, Julia Schorlemmer, Julia Schoierer, Maximilian Edlinger, Stephan Bose-O’Reilly

## Abstract

**Background:** Ongoing climate change has several indirect and direct health consequences. Children are among the most vulnerable group to suffer from these health impacts. Parents as caregivers play a particularly important role in protecting them adequately.

**Objectives:** The aim of the study was to investigate how parents perceive the health consequences of climate change. Of particular interest were their information status and already used communication channels, to make them more addressable. In addition, their risk perception and relevance estimation of the health consequences of climate change for their children were investigated.

**Methods:** Parents were able to participate anonymously in the study from March to June 2020 by means of an online questionnaire. The study sample consisted of 292 parents living in Germany (age: *M =* 42.02; *SD =* 7.73; sex: ♀ = 190; ♂ = 54) with at least one child aged 0 to 18 years. Open-ended questions and closed-ended questions with Likert scales were used. Data were analyzed descriptively. Correlations and regression analysis were performed.

**Findings:** About 75% of the respondents knew at least one health consequence of climate change. Heat related illnesses were named as the most important health consequences (74.1%). Parents obtain most of their information from the Internet (73.3%). The increase of air pollutants is estimated as the most relevant risk for the health of their children. Relevance estimation affects risk perception.

**Conclusion:** Parents are aware of the importance of being informed about the health consequences of climate change. Therefore, knowledge about the health consequences in relation to the relevance assessment must be communicated clearly and sufficiently. Information channels already used should be improved and further multipliers identified.

## Background

### Climate change as a serious health threat for children

Climate change is one of the greatest threats to human health in the 21st century^1^. According to the report of the intergovernmental panel on climate change (IPCC), any increase in global warming will affect human health, with predominantly negative consequences^2^. Health effects of climate change can be both direct (e.g. increase in extreme weather events) and indirect (e.g. increase in allergens)^3^. The system of classification shown in Figure 1 provides a basis for understanding the impact of climate change and their related hazards as well as the resulting health consequences.

**Figure 1:**
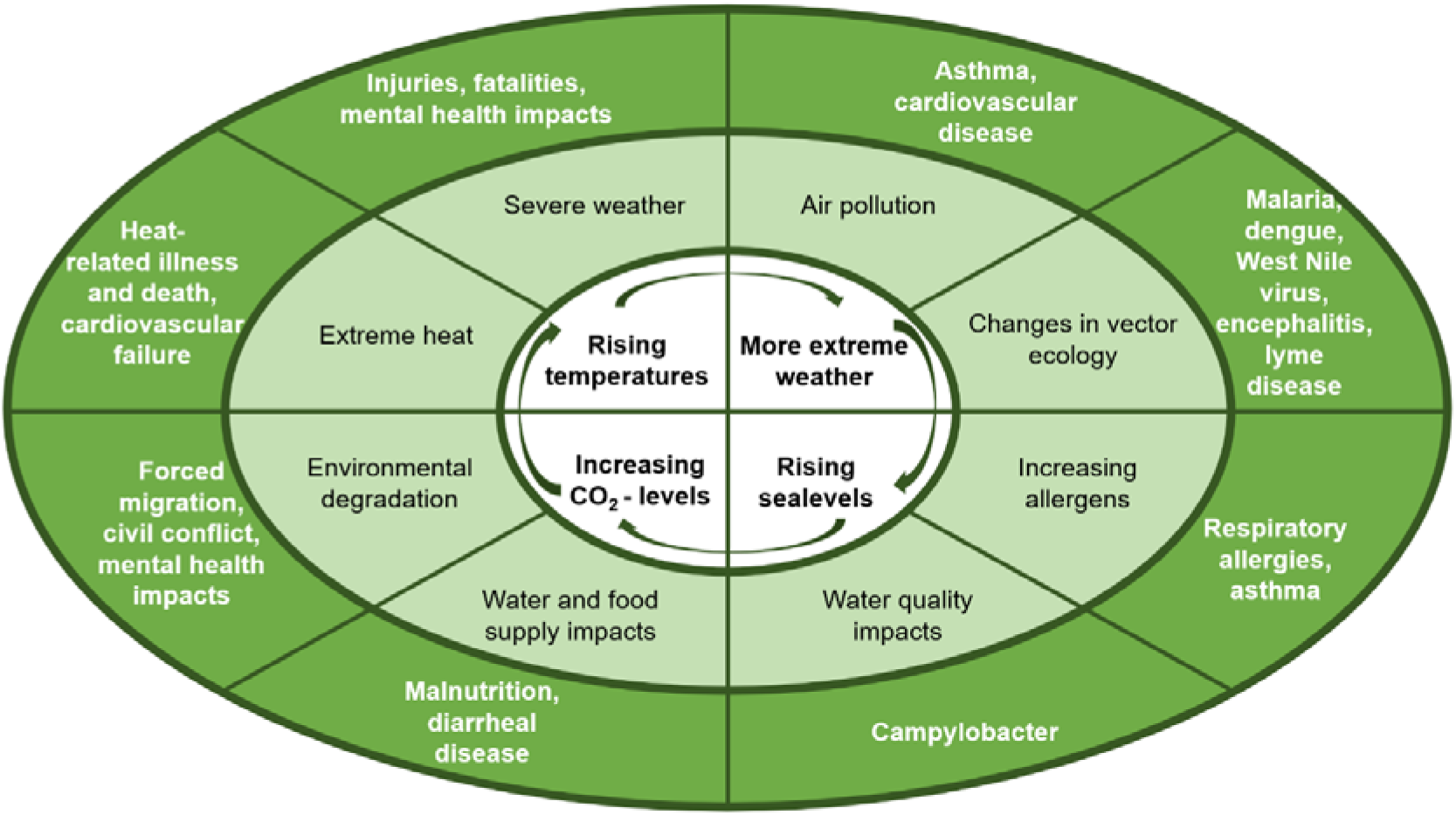
Impact of climate change, related hazards and health consequences. Adapted from National Center for Environmental Health^8^.

It is now well established that climate change can impair all population groups, however, children are considered as particularly threatened^4^. As noted by the World Health Organization (WHO) in total 88% of global burden of disease arising from climate change relate to children younger than five years^6^. They are more likely to suffer from health consequences due to climate change because they prospectively live longer, are dependent of caregivers and they are more exposed per unit body weight. Since their baseline metabolism and physiology alters from adults, children are rather able to adapt to heat^4^. The thermoregulatory capacity is not yet entirely developed in children up to four years of age, what makes them especially vulnerable to suffer from overheating during heat waves^7^.

### Preparedness and risk communication for parents

The protection of children against the expectable health consequences of climate change implements that parents are aware of the health problems, perceive a risk for their children and are willing to change their behavior. Therefore parents should be adequately prepared for the effects through campaigns of responsible institutions (e. g. for Germany the Robert-Koch-Institut^9^). Climate change appears with several factors, which complicates the communication of risks, not only for health consequences, but also for other environmental issues^10^. The communication of hazards related to climate change differs from other risk communication regarding topics like health problems and is therefore more complicated. Relevant factors are among others invisible and abstract causes, disconnection between cause and effect and self-interested behaviour^10^. An appropriate risk communication should improve knowledge and risk perception, which contributes to individual behaviour^11^. It is clear, why particular parents as caregivers should be/ get “prepared” when thinking about possibilities to protect children from health consequences of climate change. To assure an adequate risk communication, preferred communication channels and existing communication channels must be identified to reach the target group as good as possible. In general, parents seem to use a broad variation of information sources for child health advices. A study shows that parents rely on child health advices given by pediatricians more than on information provided through media such as internet, newspaper and television^12^. Especially young children and their parents have regularly contact with their pediatrician. In Germany for example a minimum of 89% of all children (age ≥ 6) have annually contact with a pediatrician in an ambulant setting^13^. Therefore, a quantitative study was conducted to investigate, which effects of climate change on human health parents already know and how relevant they consider them to be. For interest were already used information channels. It was also examined, what risks parents perceive for the health of their own child. To determine factors that contribute to parents risk perception, the effect of knowledge and relevance estimation was analyzed (Figure 2).

**Figure 2:**
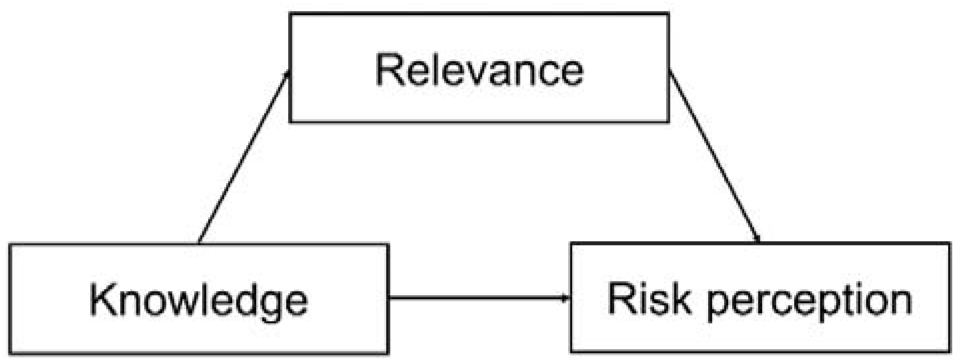
Illustration of effect of knowledge and relevance on risk perception.

## Methods

### Study procedure and design

As part of the field ‘climate change and children’s health’ an online questionnaire in German lasting maximum 10 minutes was configured using the open source software tool Lime Survey. It was first applied in a pretest version. The distribution via social networks and messengers started in March and ended in June 2020. All participants could take part anonymously on a voluntary basis and were informed about the purpose of the study.

### Study population

Participation was possible for all parents living in Germany with at least one child aged 0 to 18 years. The final study population consisted of *N* = 292 persons (*M =* 42.02; *SD =* 7.73; sex: ♀ = 190; ♂ = 54), who had most often two children (*M* = 1.91; *SD* = .88), with on average 9.02 years of age (*SD* = 6.77). In total 461 parents opened the questionnaire, 138 were excluded, because they only answered the first question (“Do you have children?”). Additionally 31 persons with the youngest child older than 18 years were not considered.

### Measures and scales

The questionnaire can be divided in the following constructs and scales in the context of climate change and health: information status, relevance estimation, risk perception and sociodemographic information. Using 4-point Likert scales (1 = “totally disagree”; 2 = “disagree”; 3 = “agree”; 4 = “totally agree”) risk perception and relevance estimation (Cronbach’s-α = 0.615) was measured. Additionally knowledge was operationalized with two items (Cronbach’s-α = 0.712) concerning information status.

To assess whether and what parents know about the negative health consequences of climate change they were asked to name the first three effects they have in mind. The given answers were then classified in the categorization of the National Center for Environmental Health (Figure 1)^8^. In order to identify relevant information channels participants should choose all information sources they already used in the context of climate change and health. Important was also to measure parents’ estimation of relevance of climate change consequences on their children’s health^3^. Risk perception was established investigating whether a person feels worried with regard to the expected effects.

### Statistical methods

Data management and analysis were carried out using the 25^th^ version of SPSS ®. Reported will be the descriptive results. Pearson’s correlation coefficient was computed. Multiple linear regression analysis as well as a mediation analysis were performed to investigate the effect of knowledge and relevance estimation on risk perception. Significance levels were set at the 5% level^14^.

### Ethic statement

The local ethics committee of the University Hospital in Munich approved the study (# 20-034). The study was orientated on the Declaration of Helsinki^15^. Since the study was fully anonymized no informed consent was needed.

## Findings

### Knowledge and information status

Considering the open question of already known consequences of climate change on health, half of those surveyed (52%) gave three answers. Altogether 75.3% named at least one health effect on human health, other answers referred to changes in the ecosystem (see Figure 3).

**Figure 3:**
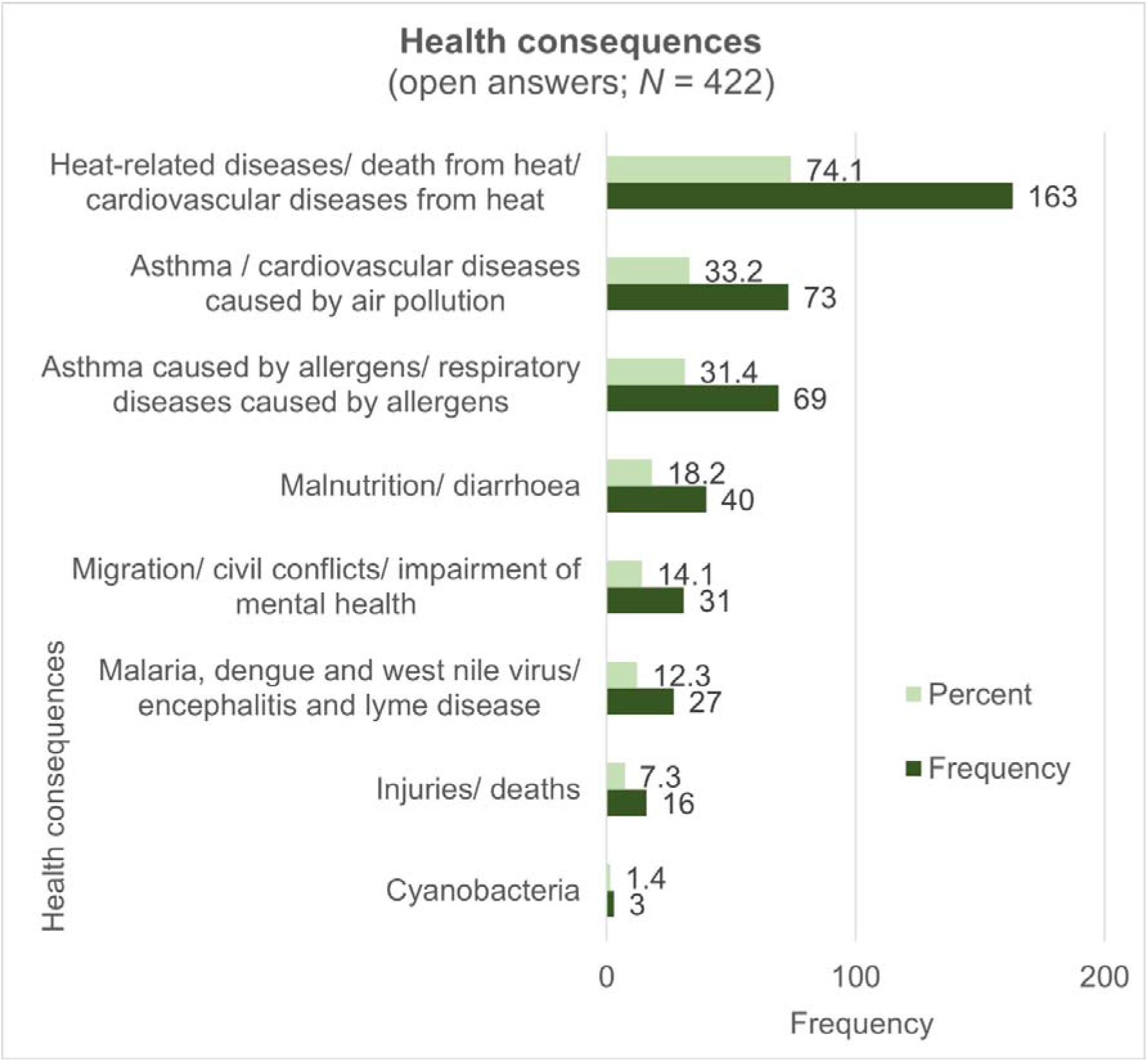
‘Which consequences of climate change on human health do you know?’ (open question, maximal 3 answers possible).

Parents were asked how informed they feel about the health consequences through climate change. It was shown that half of the parents (53%) disagreed feeling well informed (selected 1 = “totally disagree” or 2 = “disagree”) (*M* = 2.49; *SD* = 0.90). Furthermore, one question was how and where parents inform themselves. Overall 74.1% strongly agreed or agreed knowing, where to search for information (*M* = 3.0; *SD* = 0.93). When asked about the already used information sources, parents appeared to inform themselves mainly via internet and social media (73.3%). From great importance are as well print media like newspaper (46.9%) and books/magazines (43.5%) as television (42.5%). Discovering the role of pediatricians, it becomes clear that participants use them as information source only little (16.8%) (Figure 4).

**Figure 4:**
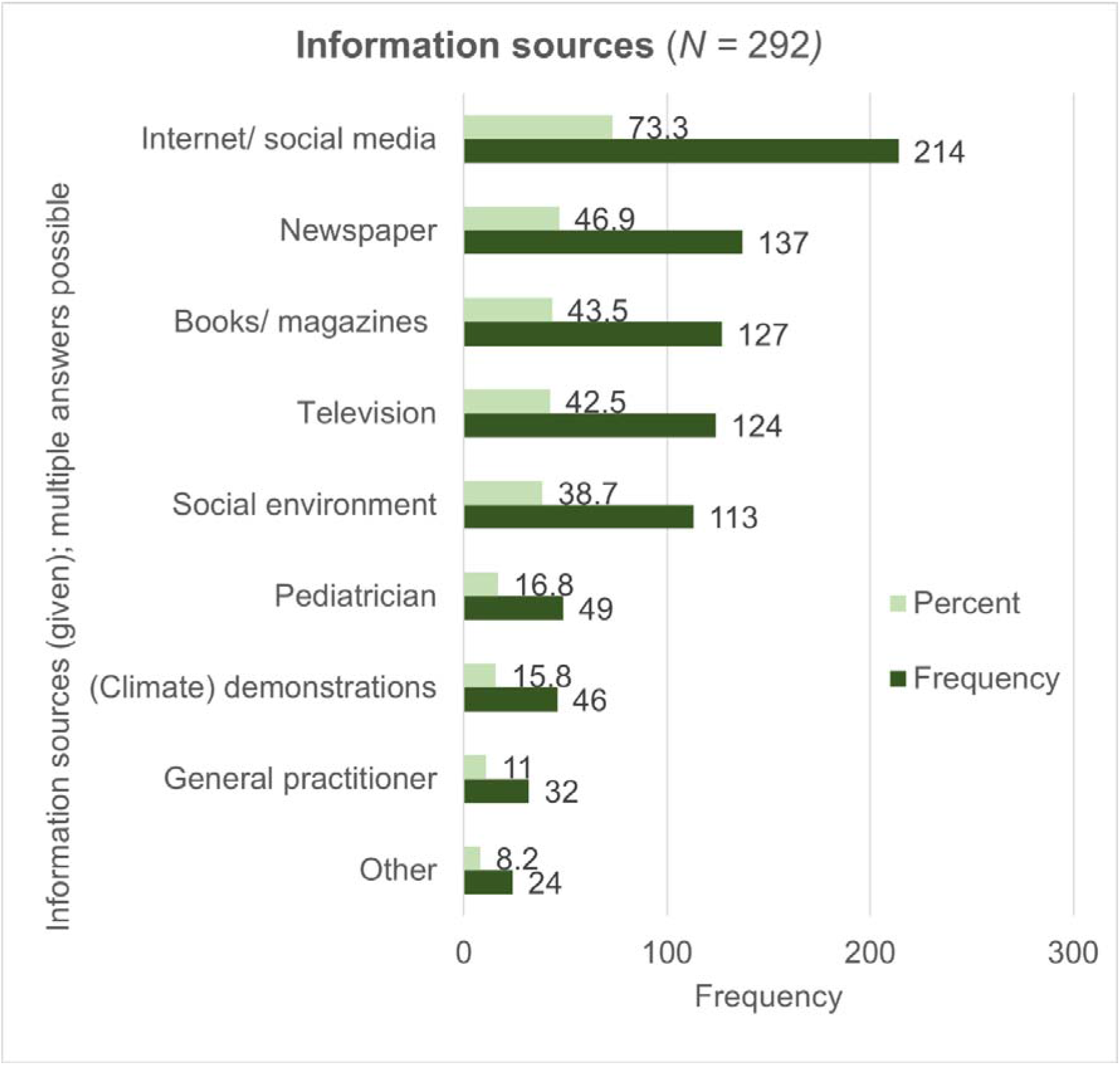
‘Where do you find information about consequences of climate change on human health’ (Information sources given, multiple answers possible).

### Relevance estimation

Estimating the relevance of consequences named in the open question, 63.6% of participants rated them as very relevant (*M* = 3.56; *SD* = 0.52). When the expectable consequences^3^ were given, it can be seen in Figure 5 that parents see the highest risk for the health of their children through the increase of air pollutants (*M* = 3.53; *SD* = 0.72), the increase of UV-radiation (*M* = 3.36; *SD* = 0.70) and the higher contamination of water with pathogens like Cyanobacteria (*M* = 3.04; *SD* = 0.90). The mean score estimating the risk for all consequences was 2.84 (*SD* = 0.44).

**Figure 5:**
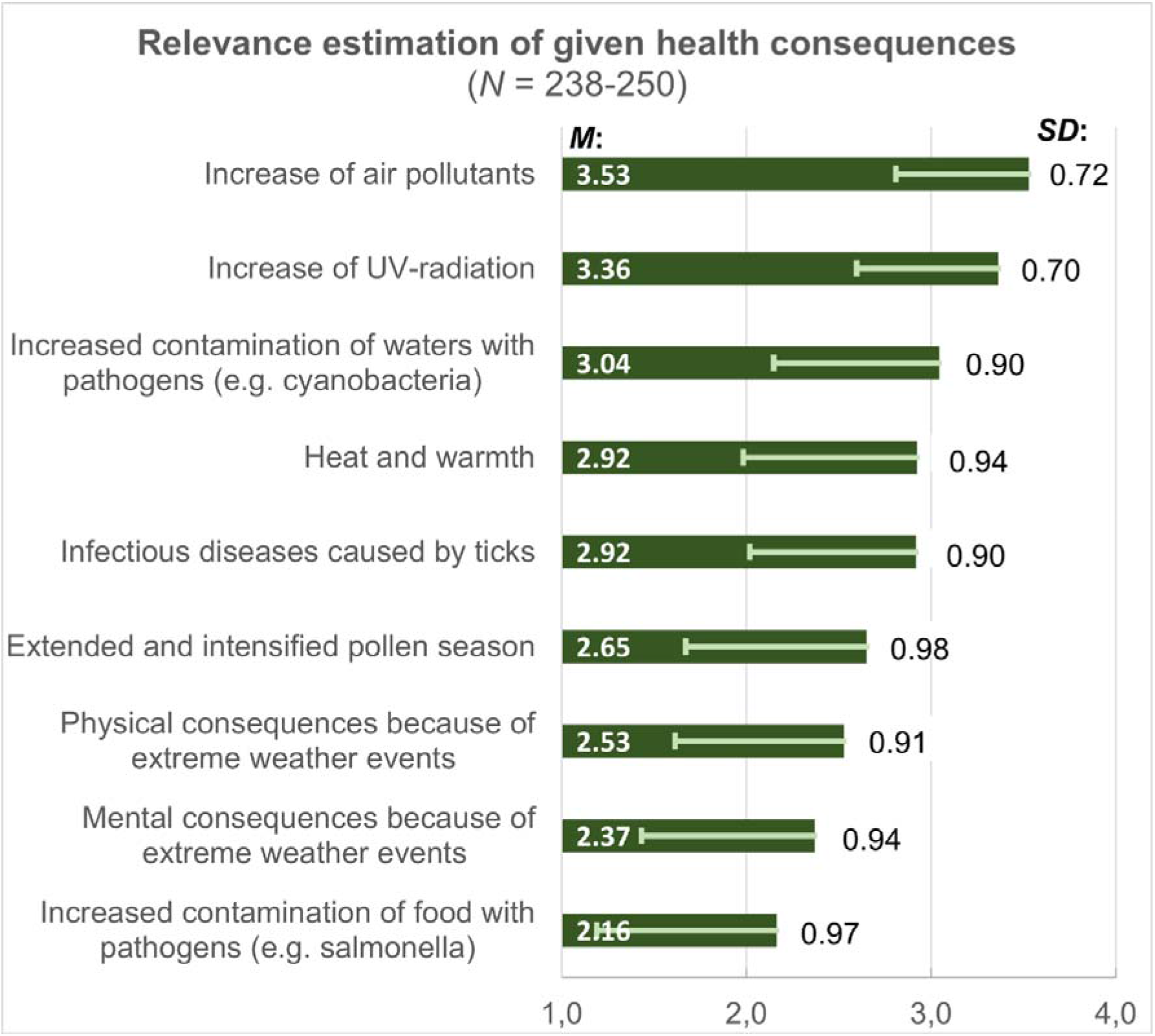
Relevance of ‘I perceive a risk for the health of my child through [given health consequences].’ 4-point-likert scale used (1 = “totally disagree”; 2 = “disagree”; 3 = “agree”; 4 = “totally agree”). Mean values and standard deviations are reported.

### Risk perception

Establishing the risk perception, all participants were asked whether climate change and its consequences worry them. The results show that the majority of all participants (73%) feels worried (*M* = 3.50; *SD* = 0.78). In addition, it was asked in which period of time the participants consider the topic of climate change and health to be important. For 89.8% the topic is already relevant. Only small proportions say that it will be important for them in five years (6.1%), ten years (2.4%) or never, because it would not concern them (1.6%).

### Relatedness within the constructs knowledge, relevance and risk perception

The following set of analysis examined whether within the constructs knowledge, operationalized through information status, relevance and risk perception exist any correlations. Table 1 provides the summary statistics for the intercorrelations among the three constructs. A medium positive correlation was found between relevance estimation and risk perception (*r* = 0.38; *p* < 0.001). Respondents with a high relevance estimation therefore perceive a higher risk.

**Table 1:** Intercorrelation matrix. Means, standard deviation and correlations of used scales are reported (1 = “totally disagree”; 2 = “disagree”; 3 = “agree”; 4 = “totally agree”).

|  | *M* | *SD* | 1 | 2 | 3 |
| --- | --- | --- | --- | --- | --- |
| 1 Knowledge | 2.74 | 0.80 |  |  |  |
| 2 Relevance | 2.84 | 0.44 | 0.10 |  |  |
| 3 Risk perception | 3.50 | 0.78 | -0.03 | **0.38\*\*** |  |
| *N* |  |  | 249 | 250 | 247 |

Note: Knowledge operationalized by scale Information status ("I know exactly where I can find information on the topic"; "I feel sufficiently well informed on the topic"). Relevance operationalized by relevance estimation of the given health consequences ("I see a risk to my child's health from [given effects of climate change on health]"). Risk perception operationalized by "Climate change and its consequences worry me".
Correlation according to Pearson, interpretation of correlation coefficient<sup>16</sup>: small effect at $|r| = 0.10$ ; medium effect at $|r| = 0.30$ ; large effect at $|r| = 0.50$ .
Bold marked are all significant values, \*. $p < 0.05$ (2-sided) significant, \*\*. $p < 0.001$ (2-sided).

In order to investigate if knowledge and relevance estimation predict risk perception in a second step regression analysis was used^14^. Gender, occupation (medicine/ health professions vs. other) and education (university degree vs. other) were considered as control variables. The obtained results are set out in Table 2. They show that relevance estimation significantly predicts risk perception. There was no evidence that knowledge has an influence on the perceived risk as (Table 2). The mediation effect of knowledge between relevance on risk perception could not be shown in the analysis.

**Table 2:**
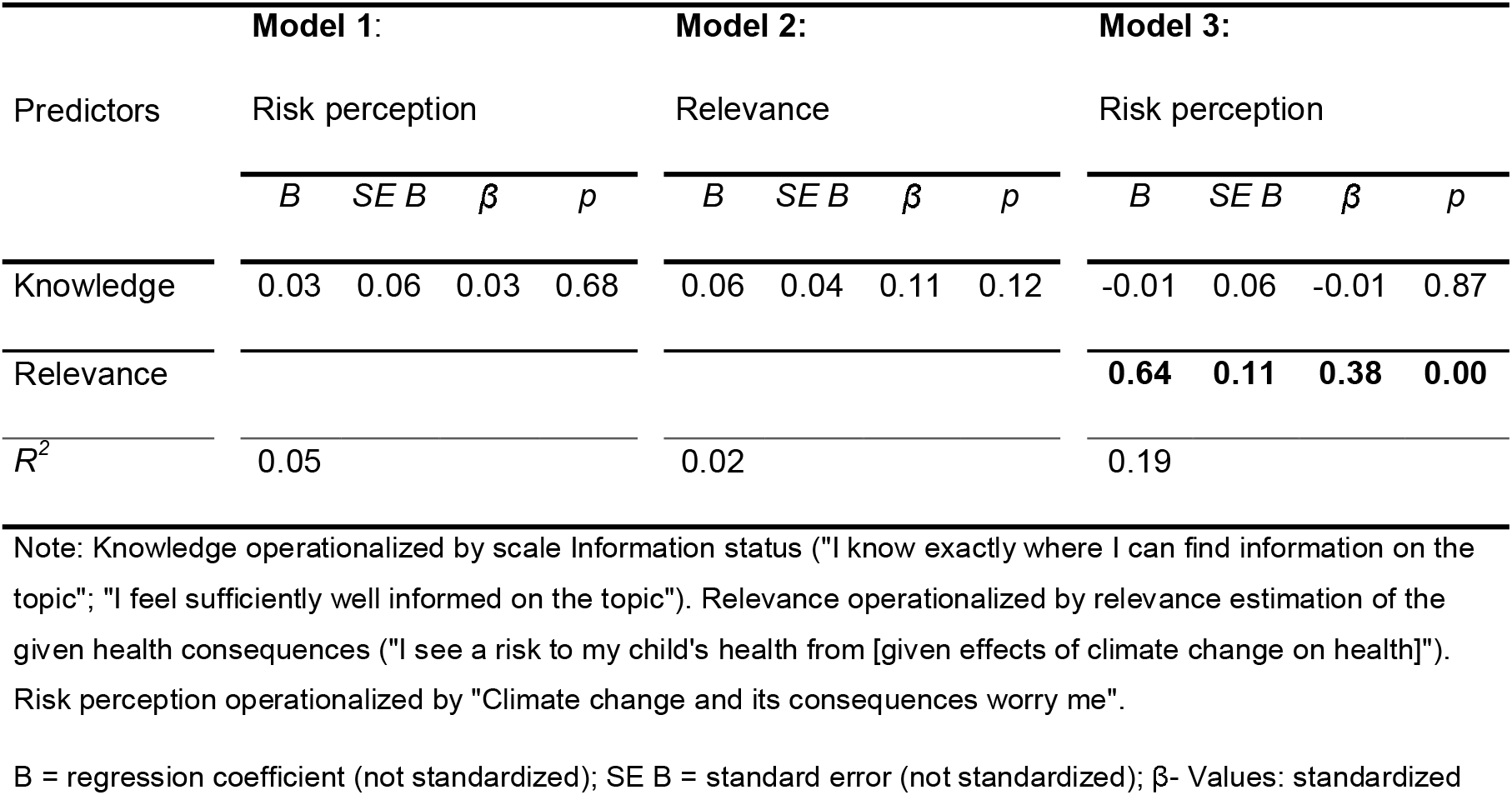
Investigation of the effect of knowledge and relevance estimation on risk perception using linear regressions. The following control variables were considered in all models: Gender, occupation (medicine/ health professions vs. other) and education (university degree vs. other).

## Discussion

In the context of climate change and children’s health, the present study was designed to determine the current information status, relevance estimation and risk perception of parents. Furthermore, it was set out with the aim of identifying relevant information sources and possible multiplicators to reach parents through risk communication more effectively. Risk perception is a relevant predictor for changing or adapting behavior, therefore this study examined antecedence for risk perception of parents.

The results of the conducted study show that the studied sample of parents know most often consequences of climate change associated with heat, when asked about possible health problems of climate change. Furthermore about one third mentions health issues because of air pollution and allergens, such as asthma. The focus gets different when asked about the perceived risk of the expectable health consequences. Then participants mostly perceived a risk for their children because of increasing air pollutants, followed by changes of UV-radiation. One explanation for the difference could be that air pollutants are in general perceived as a health risk, rather than heat-related problems. The consequences of heat are currently slightly noticeable in Germany and stay primarily abstract^3^, therefore a lower relevance for the own child’s health might be perceived. Over to thirds of participants indicate that they know where to search for information about climate change and health. But half of the parents feel uninformed. Taken together, these results suggest that the already used sources might not be fully sufficient for providing information. Parents primarily seek information via internet (/ social media), newspaper and books/ magazines. In contrast, pediatricians (and general practitioners) are rarely considered as suitable source of information.

On the question, which factors impact parents’ actions towards more prevention against health consequences of climate change, this study focused on risk perception as requirement of intentions for actions^11^. Therefore, the effect of knowledge and relevance estimation on risk perception was examined. It has been suggested that knowledge has an impact on risk perception^11^. This does not appear to be the case. The results presented in the conducted study show that knowledge alone does not seem to predict risk perception. On the other hand, relevance estimation has a statistical significant effect on risk perception. It can therefore be assumed that a person will rather bring up intentions for individual behavior and preventive actions in the context of climate change and health when relevance for the own child is recognized. The results suggest that it is not sufficient to only provide information or knowledge about climate change and health consequences, the relevance for the own child should as well be demonstrated and linked with experiences of the parents that already happen or could happen in future. Risk perception and - in a second step – intentions towards preventive actions could in that way be supported.

Our literature review revealed very little on the topic what parents already know about health consequences of climate change. A representative population survey showed that when asking about risks due to climate change, only 14.1% of all participants name health consequences^17^. For over a half (51.7%), and thus most often, natural disaster and weather problems were perceived as risks. When asked about the specific health consequences due to climate change, participants felt in average worried. It can be clearly seen that they felt especially worried about more frequent occurrence of heat waves, drought and forest fire. Equally worrying are the more frequent occurrence of storms and floods^17^. In contrast, participating parents have rated the risk of the single consequences differently (Figure 5) as shown before. For them especially air pollution an UV-radiation seem to be relevant. A study conducted in the U.S.^18^ came up with similar results compared to the one just introduced^17^, here especially open questions showed missing associations about climate change and consequences for human health^18^. This result may be explained by the fact that media articles cover the topic climate change and human health only very seldom^1^. When taking the newspaper the *People’s Daily* into account, within ten years (2008-2018) on average about 2500 articles each year addressed climate change, from which only 14 articles a year focused also on human health^1^. Mass media might in some cases have an impact on a person’s risk perception, whereas interpersonal communication channels may be effective as well^19^.

Compared to this, the ongoing SARS-CoV-2 pandemic has a great media coverage. An annual study conducted between February and April investigated the general risk perception of persons living in Hamburg, Germany^20^. It has been demonstrated that the personal risk perception has decreased in 2020, which is different to the prior years since 2014. Simultaneously the new coronavirus has been perceived as a major problem, which is ranked higher than environmental problems. During this time period, the prevention measures to combat the SARS-CoV-2-virus have had a strong impact on the daily life of participants. As a result, the risk perception regarding the new coronavirus might have been higher compared to the more abstract consequences of climate change^20^.

### Limitations

The results of the conducted study mostly account for a selective group of parents. Most of participants were female (78%) and born in Germany (94%). Furthermore, a high level of education was predominant and a large number of participants worked in the health sector/ had medical professions (26%). Therefore, external validity might be low and some vulnerable groups might not be fully reached. On the one hand, it was shown that the studied sample seems to be well informed, but on the other hand, there still are insecurities and discrepancies which should be addressed. Beginning with improving this knowledge-gap for well educated parents, other groups might be reached as well.

### Future Research

Further research should be undertaken to investigate how parents want to be informed/could be better informed about health and climate change in generals and about the direct impact on their children’s health in the future. The conducted study did not demonstrate in which way already used information channels should be improved for a better risk communication. It remains unclear, which further communication channels and/ or multiplicators (for example pediatricians) might be desired and most effective. Developments of suitable risk communication about the expectable health consequences due to climate change and adaption strategies based on the existing knowledge of parents are therefore recommended. Since the important meaning of relevance estimation has been shown, further research should also focus on possibilities to link relevance estimation with knowledge. Therefore the meaning of the health consequences of climate change for the own child now and in the future has to be rather demonstrated in a way that increases the risk perception of parents and leads to behavior change as well.

## Conclusions

The aim of this study was to gain insights into what parents already know about the health consequences of climate change and what they do not yet sufficiently know. The parents are primarily aware of heat-related illnesses, asthma due to air pollution or allergens. They see a risk for their children primarily from an increase in air pollutants and UV-radiation. These health problems are relevant to parents now, and not just in the future. Overall, the study reinforces the idea that parents are willing to inform themselves about the health consequences of climate change (for their children). The results, combined with future research on the health dimensions of climate change, will therefore be of interest to policymakers who provide adaption strategies. Some questions remain to be answered. The provision and presentation of information about the health consequences of climate change and related adaptation strategies should be more relevant to the individual child in order to increase parents’ perception of risk.

## Data Availability

Data are available on reasonable request.

## Notes

**Funding sources**: Funded by the Federal Ministry for the Environment, Nature Conservation and Nuclear Safety (BMU) on the basis of a resolution of the German Bundestag (project number: 67DAS213)

**Conflict of interest statement for all authors** The authors declare that there is no conflict of interest regarding the publication of this paper.

### Competing Interest Statement

The authors have declared no competing interest.

### Clinical Trial

The research is not declared as clinical trial, however the local ethics committee of the University Hospital in Munich approved the study (# 20-034).

### Funding Statement

Funded by the Federal Ministry for the Environment, Nature Conservation and Nuclear Safety (BMU) on the basis of a resolution of the German Bundestag (project number: 67DAS213).

### Author Declarations

The local ethics committee of the University Hospital in Munich approved the study (# 20-034).

